# Bidirectional two-sample Mendelian randomization study of differential white blood cell count and schizophrenia

**DOI:** 10.1101/2023.05.19.23290200

**Authors:** Perry BM Leung, Zipeng Liu, Yuanxin Zhong, Marta Di Forti, Robin M Murray, Hon-Cheong So, Pak C Sham, Simon SY Lui

## Abstract

**Background:** Schizophrenia and white blood cell count (WBC) are both complex and polygenic disease/traits. Previous evidence suggested that increased WBC is associated with higher all-cause mortality, and other evidence found elevated WBC in first-episode psychosis and chronic schizophrenia patients. However, prior observational findings may be confounded by antipsychotic exposures and their effects on WBC. Mendelian randomization (MR) is a useful method to examine the directional causal relationship between schizophrenia and WBC

**Methods:** We performed a two-sample MR using summary statistics of the Psychiatric Genomics Consortium Schizophrenia Workgroup (N=130,644) and the Blood Cell Consortium (N=563,085). The MR methods included inverse variance weighted, ME Egger, weighted median, and MR-PRESSO, contamination mixture, and a novel approach called mixture model reciprocal causal inference (MRCI). False discovery rate was employed to correct for multiple testing.

**Results:** After correcting for horizontal pleiotropy, the MRCI method demonstrated that elevated lymphocyte count (causal effects at the liability scale=0.077; FDR adjusted p-value=0.026) and eosinophil count (causal effects at the liability scale=0.048; FDR adjusted p-value=0.026) may cause schizophrenia. The contamination mixture method showed that schizophrenia may lead to elevated neutrophil count (beta=0.011 in unit of standard deviation of mean absolute neutrophil count; FDR adjusted p-value=0.045) and reduction of eosinophil count (beta=-0.013 in unit of standard deviation of mean absolute eosinophil count; FDR adjusted p-value=0.045). Some further significant findings had been identified by conventional MR approaches and MR-PRESSO, but we interpreted those with cautious due to substantial heterogeneity and plausible pleiotropic effects identified.

**Conclusion:** This MR study provided evidence that schizophrenia has causal relationships with altered differential WBC. Our findings support the role of WBC in influencing schizophrenia risk, and may concur with the hypothesis of neuroinflammation in schizophrenia.

## 1. Introduction

Schizophrenia, which affects 23.6 million people globally, is causing high disability and substantial caregiver burden (GBD 2019 Mental Disorders Collaborators, 2022). The pathological mechanisms underlying schizophrenia remain largely unknown, but some hypotheses implicate the immune system (Vidal & Pacheco, 2020). Ermakov et al. (2022) summarized these including the general inflammation hypothesis, macrophage-T-cell hypothesis, autoantibody hypothesis, and microglia hypothesis.

Notably, above one-third of people with schizophrenia show abnormalities of immune biomarkers, such as abnormal levels of C-reactive protein, and dysregulations of cytokines, chemokines and microbiota (Gibney & Drexhage, 2013; Ermakov et al., 2022; Hartwig et al., 2017; Van Kestere et al., 2017). Elevated circulating levels of proinflammatory cytokines have been observed prior to onset of schizophrenia (Williams et al., 2022). Connections between complement system within the immune system via the C4 gene and development of schizophrenia have been reported (Sekar et al., 2016). Besides, epidemiological evidence has also revealed strong associations of prenatal and childhood infections with schizophrenia (Davies et al., 2020; Cheslack-Postava & Brown, 2022). In addition, Tylee et al. (2018) reported genome-wide genetic correlation between schizophrenia and various immune-related disorders, such as Crohn’s disease, primary biliary cirrhosis, systemic lupus erythematosus, and ulcerative colitis.

Differential white blood cells, which include lymphocytes, basophils, eosinophils, monocytes, and neutrophils, are indispensable to innate and adaptive immunity. Neutrophils are the most abundant and serve as the front line of innate immunity. Functions of neutrophils include damaging pathogens, producing cytokines, facilitating activation of macrophages, regulating signals for proliferation, and activations of B and T lymphocytes to activate adaptive immunity. Lymphocytes, subclassified into B cells, T cells, natural killer cells, and innate lymphoid cells, are pivotal in producing antibodies and regulating adaptive immune responses (Gasteiger & Rudensky, 2014). Monocytes can derive both pro-and anti-inflammatory macrophages under different microenvironments, as well as dendritic cells at inflammatory sites. Pro-inflammatory monocytes release multiple chemokines aiming to removing the invading pathogens, while anti-inflammatory monocytes negatively regulate proinflammatory cytokines and facilitate resolution of inflammation (Austermann et al., 2022). Eosinophils are part of the innate immune system with essential roles in bridging innate and adaptive immunity. It can affect the immune microenvironment, cellular recruitment as well as tissue homeostasis, repair and remodelling (Shamri et al., 2011). Lastly, basophils have the least proportion in blood but may act as mediators in allergic reactions, which carry critical roles in immunomodulation and are involved in many infectious, allergic and autoimmune diseases (Zhang et al., 2021).

Given the interconnection between levels of peripheral white blood cell count (WBC) and immune response, they are commonly-used as laboratory-indicators for inflammatory, immune, allergic and hematologic diseases (Jain et al., 2017). Elevated WBC are associated with higher all-cause mortality, in particular related to respiratory, cardiac and metabolic disorders (Chan et al., 2022; Kim et al, 2013; Shim et al., 2006). Although WBC fluctuates with time to adapt to environmental pathogen exposure (Hung et al., 2021), these markers also show ‘trait-like’ properties, such as persistently elevated WBC in physically-active people (Farhangi et al, 2013). The levels of WBC are heritable complex traits, following a polygenic model (Whitfield et al., 1985; Evans et al., 1999). The heritability of lymphocyte, monocyte, and neutrophil count were estimated at 58%, 58%, and 41% respectively in European samples (Lin et al., 2016; Lin et al., 2017).

Empirical evidence supports an association between schizophrenia and elevated WBC. Several meta-analyses found people with first-episode psychosis and chronic schizophrenia had elevated WBC (Miller et al., 2013; Jackson & Miller, 2020; Mazza et al., 2020). Elevated neutrophil/lymphocyte ratio (NLR) was found to be consistently elevated in people with first-episode psychosis, and chronic schizophrenia (Karageorgiou et al., 2019). A longitudinal study which followed a cohort of first-episode psychosis reported that NLR was higher in non-remitted than in remitted schizophrenia (Bioque et al., 2022). Clozapine has well-known effects in altering WBC in people with schizophrenia, including rare and potentially fatal clozapine-induced neutropenia, as well as more common and benign leukocytosis (Paribello et al., 2021). Non-clozapine antipsychotic medications, such as olanzapine and risperidone, have also been reported to alter WBC (Alvarez-Herrera et al., 2020; Rettenbacher et al., 2010). Findings regarding whether antipsychotic medications can alter NLR remain inconsistent (Zhou et al., 2020; Dawidowski et al., 2022; Bioque et al., 2022).

One method to examine the causal relationship between traits is Mendelian Randomization (MR). MR is a statistical approach to evaluate causal relevance of biomarkers (Holmes et al., 2015). It mimics randomization by using generic variants as ‘instrumental variables’ to estimate the causal effect of an exposure on an outcome of interest. This approach substantially reduces the risk of confounding bias typically encountered in case-control and cohort studies, based on the assumption that genetic variants are randomly allocated during meiosis, and subsequently independent from many confounding environmental factors and reverse causality that hamper the validity of causal linkage in observational studies. The allelic variation in genes is also independent of the disease development and progression within individuals.

There is a recent study attempted to explore the association between schizophrenia and WBC (Gao et al., 2023). Nevertheless, this prior study had shortcomings. First, the data they used for MR were not the most up-to-date, as the Psychiatric Genomics Consortium published the “third wave” data in 2022 (Trubetskoy et al., 2022), and the Blood Cell Consortium had further combined UK Biobank with other cohorts to provide the Phase 2 meta-analytic data (Vuckovic et al., 2020, Chen et al., 2021; Lettre, 2023). Moreover, several novel MR approaches have been developed to address horizontal pleiotropy and to provide outliers-, and pleiotropic-robust estimates. Different from Gao et al. (2023)’s study, we used the contamination mixture method developed in 2020 (Burgess et al., 2020), and the Mixture model Reciprocal Causation Inference (MRCI) method that our team had developed very recently (Liu et al., 2023).

This MR study aimed to explore the causal relationship between schizophrenia and WBC. Furthermore, we employed two-sample (rather than one-sample) MR to minimize the risk of biases caused by winners’ curse and weak instruments (Lawlor, 2016). To our knowledge, this bidirectional two-sample MR study is the first to utilize the most updated GWAS data provided by two large scale consortiums, namely the Psychiatric Genomics Consortium wave 3 and the Blood Cell Consortium Phase 2.

## 2. Methods

Our two-sample MR used two independent datasets as the exposure and outcome, both sensitive to causal effects of mediators and corrected for measurement errors (Carter et al., 2019). The three essential assumptions to uphold unbiased estimates of MR are (1) the genetic instruments are associated with exposure; (2) the genetic instruments are associated with the outcome only through the chosen exposure (i.e. no pleiotropic effects); and (3) the genetic instruments are independent of other factors which could influence the outcome (Boef et al., 2015).

### 2.1 Data source

#### 2.1.1. Psychiatric Genomics Consortium (PGC) schizophrenia genome-wide association study (GWAS)

We used the summary statistics gathered from the most up-to-date, third wave PGC schizophrenia GWAS core dataset, which contained 53,386 schizophrenia cases and 77,258 controls from European ancestry. Details of this dataset have been described elsewhere (Trubetskoy et al., 2022).

#### 2.1.2. Blood Cell Consortium

We used the summary statistics of differential WBC gathered from the European working groups Blood Cell Consortium Phase 2 (BCX2), through the IEU Open GWAS Project, which contained 26 discovery cohorts of European ancestry additional to UK Biobank, which total 563,946 subjects. The log10-transformed absolute count, in units of (×10^9^ cells/L), were reported after multiplying relative count with the total WBC count from the original cohorts. Detailed descriptions of each cohort and data management have been described elsewhere (Vuckovic et al., 2020, Chen et al., 2021; Lettre, 2023).

### 2.2. Statistical analysis

We first retrieved the single nucleotide polymorphisms (SNPs) from the abovementioned two GWAS meta-analysis study data, and selected those SNPs which showed a p-value smaller than the GWAS significant threshold (i.e. p-value < 5e-8). SNPs were then clumped based on the ‘TwoSampleMR’ package in statistical software R, with the default linkage disequilibrium threshold r^2^< 0.001, distance > 10000kb, and 1000 genomes European reference panel. Palindromic and ambiguous SNPs were further removed after harmonising the data between exposure and outcome datasets (i.e. the default option in TwoSampleMR R package). F-statistics were calculated to assess instrument strength. By a general rule of thumb, an F-statistic of >10 is regarded as a lack of evidence for weak instruments which (if present) could cause biases towards the null (Wootton et al., 2018).

We comprehensively employed different MR methods in this study, which were summarized in Table 1, because each has its own assumptions, strengths and limitations (Bowden et al., 2016; Burgess & Thompson, 2017; Verbanck et al., 2018; Slob & Burgess, 2020; Liu et al.,2023). In brief, random-effect inverse variance weighted (IVW) provides unbiased estimates when all three MR assumptions are clearly satisfied. Weighted median is robust to outliers when the majority of genetic variants are valid instruments (Slob & Burgess, 2020). Horizontal pleiotropy refers to existence of pleiotropic SNPs which have influences on the outcome other than through the exposure; this will violate the exclusion restriction assumption of MR and undermine the validity of the estimates. When horizontal pleiotropies are present, other MR methods should be considered (Slob & Burgess, 2020). For instance, MR-Egger relaxes the assumption of exclusion restriction by allowing the strength of instruments to be independent from the direct effect (InSIDE) assumption. InSIDE states that genetic variants-exposure pathways should be independent from the genetic variants-outcome pathways (Bowden et al., 2015). Furthermore, MR-PRESSO is an extended approach of IVW as it performs IVW analysis after removing the outliers. MR-PRESSO detects horizontal pleiotropy using the MR-PRESSO global test, and determines outliers using the MR-PRESSO distortion test, and then yield ‘outlier-adjusted causal estimates’ (Wootton et al., 2018). Nonetheless, MR-PRESSO is prone to Type 1 error, if a few invalid instruments are retained in the sample after removing the outliers (Slob & Burgess, 2020). Contamination mixture MR is a more conservative method, compared with MR-PRESSO and other outliers-removed approaches, because it assumes plurality valid and can give close to nominal rates of Type 1 error up to 40% invalid genetic variants, while maintaining reasonable power to detect causal effect (Burgess et al., 2020).

**Table 1:** The assumptions, strengths and weaknesses of selected MR methods.

| **Analysis method** | **Consistency assumption** | **Strengths** | **Weaknesses** |
| --- | --- | --- | --- |
| Random effect inverse variance weighted (IVW) | Average pleiotropic effect must be zero | Consistent and efficient estimate when MR assumptions are satisfied | All variants have to be valid IVs which is also known as 0% breakdown point |
| Weighted median | Majority (>50%) of genetic variants are valid IVs | Robust to outliers as the method focus on the median values only | Less efficient than IVW |
| MR-Egger | InSIDE (Instrument Strength Independent of Direct Effect) | The InSIDE assumption is less stringent compared with IVW | The InSIDE assumption is sometimes not plausible. This method is also highly sensitive to outliers |
| MR-PRESSO | Majority (>50%) of genetic variants are valid IVs | Outlier robust and efficient with valid IVs | High false positive rate with several invalid IVs |
| Contamination mixture | Plurality valid | Robust to outliers | Sensitive to variance parameters and addition/removal of genetic variants |
| Mixture model Reciprocal Causation Inference (MRCI) | Bivariate normality of standardized marginal effects in SNPs | Provide robust estimates even if most SNPs were pleiotropic | The robust sandwich standard errors are conservative |

We also employed a novel MR method, namely the Mixture model Reciprocal Causation Inference (MRCI) (Liu et al., 2023). When dealing with both independent and correlated pleiotropy, MRCI utilizes all the SNPs gathered from GWAS summary statistics to provide unbiased estimates with non-inflated Type 1 error. Independent pleiotropy occurs when a SNP contributes to at least two traits with independent mechanisms. Correlated pleiotropy, on the other hand, refers to SNPs that contribute to multiple traits with shared mechanisms. The MRCI method is unique in utilizing all the available SNPs and modelling the pleiotropy explicitly by classifying SNPs into phenotype-specific, pleiotropic, or not contributing towards the specified phenotypes, based on marginal effect of each SNP. As such, MRCI provides robust estimates even when the majority of the causal variants are pleiotropic.

Moreover, MRCI can operate under different (sometimes contradicting) assumptions (e.g., none of the variants are pleiotropic; all of the variants are pleiotropic; as well as situations where phenotype-specific variants could not be identified). The only required parameters to use MRCI were prevalence of schizophrenia which was set at 0.35% (World Health Organization, 2022) and the case proportion in our data which was 40.86%. MRCI outputs are the units of standard deviation increase for continuous variables (i.e. WBC), and heritability in liability scale for binary variable (i.e. schizophrenia).

Two-sided p-value was used for each analysis. False discovery rate (FDR) adjusted p-value were used as the conservative estimates to address the issue of multiple hypothesis testing. False Discovery rate (FDR) correction p-value controlled the expected proportion of falsely rejected hypotheses in multiple testing (Benjamini & Hochberg, 1995). In this study, an FDR < 0.05 would be deemed as the statistically significant threshold to reject null hypothesis, and FDR < 0.1 as suggestive associations. Besides, Cochran’s Q statistic and heterogeneity statistic I^2^ were computed in different harmonized datasets to test for the validity of the MR assumptions. Scatter plot, forest plot, funnel plot, and leave-one-out analysis were generated to aid the detection of outliers (see Supplementary Figures 1-8). Furthermore, power calculation was performed using an online power calculator designed for MR studies, and suggested that 563,085 participants from the Blood Cell Consortium exceeded the minimum sample size to achieve 80% power (i.e. 436,100) to detect exposure R^2^ and causal effect size as low as 0.02 and 0.03 standard deviation change respectively. On the other hand, 161,405 individuals from PGC with 80% statistical power can detect as little as 0.033 exposure R^2^ and 1.08 odds ratio per standard deviation change in outcomes (Burgess, 2023).

All the MR analyses conducted in this study were performed using ‘TwoSampleMR’ packages in the R version 4.1.0. MRCI algorithm is publicly available on GitHub (Liu et al., 2023).

## 3. Results

We found 20,457 SNPs from PGC-SZ smaller than the GWAS threshold and retained 146 of them after clumping and harmonising. F-statistics >10 was reaffirmed (refer to Supplementary Table 1). Results from all the conventional MR methods were summarized in Table 2. Using random effect IVW analysis, our results showed people with schizophrenia would have elevated peripheral lymphocyte count (beta=0.026; FDR adjusted p-val=0.008), and elevated monocyte count (beta=0.016; FDR adjusted p-val=0.067), as well as plausible higher neutrophil count (beta=0.014; FDR adjusted p-val=0.099) as suggestive associations. Moreover, the more conservative estimate from the weighted median method further illustrated schizophrenia could yield increased lymphocyte count (beta=0.013; FDR adjusted p-val=0.049). On the other hand, hundreds of SNPs from different differential WBC were included after clumping and the F statistics were calculated (refer to Supplementary Table 2). The same set of conventional MR methods were used in studying how WBC might lead to schizophrenia. The results were summarized and reported in Table 3. Notably, IVW (OR=1.100; FDR adjusted p-val=0.021) and weighted median method (OR=1.136; FDR adjusted p-val=0.006) showed that lymphocyte count are risk factors of schizophrenia, suggesting a bidirectional relationship between schizophrenia and lymphocyte count.

**Table 2:** MR results of schizophrenia on differential WBC count (β-coefficient are interpreted in units of standard deviation increase of absolute differential white blood cell count)

| Outcomes | MR methods | No. of SNP | Beta | P-value | FDR adjusted p-value |
| --- | --- | --- | --- | --- | --- |
| Basophil | IVW | 146 | 0.008 | 0.152 | 0.190 |
| Basophil | MR Egger | 146 | 0.078 | 0.0002** | 0.0003** |
| Basophil | Weighted median | 146 | 0.0009 | 0.858 | 0.895 |
| Basophil | MR-PRESSO | 139 | 0.003 | 0.512 | 0.640 |
| Basophil | Contamination mixture | 110 | -0.007 | 0.187 | 0.312 |
| Eosinophil | IVW | 146 | 0.010 | 0.206 | 0.206 |
| Eosinophil | MR Egger | 146 | 0.086 | 0.006** | 0.006** |
| Eosinophil | Weighted median | 146 | -0.008 | 0.108 | 0.180 |
| Eosinophil | MR-PRESSO | 129 | -0.0006 | 0.885 | 0.885 |
| Eosinophil | Contamination mixture | 102 | -0.013 | 0.016** | 0.045** |
| Lymphocyte | IVW | 146 | 0.026 | 0.002** | 0.008** |
| Lymphocyte | MR Egger | 146 | 0.161 | 4.7e-7** | 2.3e-6** |
| Lymphocyte | Weighted median | 146 | 0.013 | 0.010** | 0.049** |
| Lymphocyte | MR-PRESSO | 131 | 0.014 | 0.002** | 0.010** |
| Lymphocyte | Contamination mixture | 93 | 0.006 | 0.299 | 0.374 |
| Monocyte | IVW | 146 | 0.016 | 0.027** | 0.067* |
| Monocyte | MR Egger | 146 | 0.116 | 3.7e-5** | 9.3e-5** |
| Monocyte | Weighted median | 146 | -0.0006 | 0.895 | 0.895 |
| Monocyte | MR-PRESSO | 129 | 0.009 | 0.027** | 0.045** |
| Monocyte | Contamination mixture | 99 | 0.001 | 0.819 | 1.000 |
| Neutrophil | IVW | 146 | 0.014 | 0.059* | 0.099* |
| Neutrophil | MR Egger | 146 | 0.100 | 0.0007** | 0.0008** |
| Neutrophil | Weighted median | 146 | 0.010 | 0.041** | 0.103 |
| Neutrophil | MR-PRESSO | 122 | 0.011 | 0.011** | 0.028** |
| Neutrophil | Contamination mixture | 99 | 0.011 | 0.018** | 0.045** |

\*p-value<0.1 ,\*\*p-value<0.05

**Table 3:** MR results of differential WBC count on schizophrenia (β-coefficient are interpreted in units of log odds)

| Exposures | MR methods | No. of SNP | Beta | Odds ratio (OR) | P-value | FDR adjusted p-value |
| --- | --- | --- | --- | --- | --- | --- |
| Basophil | IVW | 193 | 0.107 | 1.113 | 0.047** | 0.117 |
| Basophil | MR Egger | 193 | 0.065 | 1.067 | 0.545 | 0.545 |
| Basophil | Weighted median | 193 | 0.082 | 1.086 | 0.159 | 0.264 |
| Basophil | MR-PRESSO | 186 | 0.052 | 1.053 | 0.255 | 0.638 |
| Basophil | Contamination mixture | 137 | 0.009 | 1.009 | 0.797 | 0.797 |
| Eosinophil | IVW | 428 | 0.040 | 1.041 | 0.191 | 0.319 |
| Eosinophil | MR Egger | 428 | 0.078 | 1.081 | 0.188 | 0.235 |
| Eosinophil | Weighted median | 428 | 0.064 | 1.066 | 0.070* | 0.174 |
| Eosinophil | MR-PRESSO | 417 | 0.005 | 1.005 | 0.865 | 0.912 |
| Eosinophil | Contamination mixture | 306 | 0.012 | 1.012 | 0.685 | 0.797 |
| Lymphocyte | IVW | 484 | 0.095 | 1.100 | 0.004** | 0.021** |
| Lymphocyte | MR Egger | 484 | 0.172 | 1.188 | 0.013** | 0.065* |
| Lymphocyte | Weighted median | 484 | 0.127 | 1.136 | 0.001** | 0.006** |
| Lymphocyte | MR-PRESSO | 462 | 0.045 | 1.046 | 0.125 | 0.625 |
| Lymphocyte | Contamination mixture | 336 | -0.009 | 0.991 | 0.773 | 0.797 |
| Monocyte | IVW | 492 | 0.017 | 1.017 | 0.535 | 0.535 |
| Monocyte | MR Egger | 492 | 0.081 | 1.084 | 0.083* | 0.204 |
| Monocyte | Weighted median | 492 | 0.003 | 1.003 | 0.922 | 0.997 |
| Monocyte | MR-PRESSO | 470 | 0.002 | 1.002 | 0.912 | 0.912 |
| Monocyte | Contamination mixture | 351 | 0.025 | 1.025 | 0.194 | 0.797 |
| Neutrophil | IVW | 412 | 0.032 | 1.033 | 0.410 | 0.512 |
| Neutrophil | MR Egger | 412 | 0.125 | 1.133 | 0.122 | 0.204 |
| Neutrophil | Weighted median | 412 | -0.0002 | 1.000 | 0.997 | 0.997 |
| Neutrophil | MR-PRESSO | 399 | 0.011 | 1.011 | 0.742 | 0.912 |
| Neutrophil | Contamination mixture | 282 | 0.017 | 1.017 | 0.620 | 0.797 |

\*p-value<0.1 , \*\*p-value<0.05

Nevertheless, Cochran’s Q and I^2^ statistics (see Supplementary Tables 1 & 2) indicated that there is substantial heterogeneity of the included genetic instruments, which could bias the results. Scatter plots and leave-one-out analysis (see Supplementary Figures 1-8) further confirmed the existence of a dominant SNP rs13195636. Rs13195636 was the SNP with the smallest p-value in the schizophrenia GWAS after clumping, and it also contributed heavily to the significant results on various differential WBC. This outlier SNP could contribute to horizontal pleiotropy that can bias the results from conventional MR methods. The MR-Egger intercept test and the MR-PRESSO global test also showed significant evidence of horizontal pleiotropy. As a result, the exclusion restriction assumption required by conventional MR approaches were violated which correspondingly leaded to biased estimates. Therefore, further MR methods were used to obtain outlier-robust estimates.

MR-PRESSO and contamination mixture methods both derived outlier-robust estimates by removing the outliers. Using MR-PRESSO method, we removed outlier and pleiotropic SNPs such as rs13195636, and the results suggested that schizophrenia might increase lymphocyte count (beta=0.014; FDR adjusted p-val=0.010), monocyte count (beta=0.009; FDR adjusted p-val=0.045) and neutrophil count (beta=0.011; FDR adjusted p-val=0.028). Nevertheless, only a handful of most significant SNPs were excluded using MR-PRESSO, and the Cochrane Q’s values indicated considerable heterogeneity remained (see Supplementary Tables 1 & 2). Therefore, these results should be interpreted with caution. Cochrane Q’s values showed that contamination mixture method reduced the heterogeneity of included SNPs substantially (see Supplementary Table 1), and the results suggested that schizophrenia might increase neutrophil count (beta=0.011; FDR adjusted p-val=0.045). Interestingly, contamination mixture method suggested that schizophrenia reduces eosinophil count (beta=-0.013, FDR adjusted p-val=0.045), which we did not observed from other methods. Conversely, MR-PRESSO and contamination mixture method did not suggest causal relationship from WBC to schizophrenia.

Furthermore, Table 4 summarises the results of the novel MRCI method, which suggested that both eosinophil count (causal effects at the liability scale=0.048; FDR adjusted p-val=0.010) and lymphocyte count (causal effects at the liability scale=0.077; FDR adjusted p-val=0.010) may causally link to development of schizophrenia after modelling the SNPs that contribute to pleiotropic effects to both schizophrenia and WBC. Supplementary figures 9-13 reported the detailed results of MRCI methods on schizophrenia and differential WBC, which included all the reciprocal causal estimates from each model (Panel A), weights of different models that the algorithm took into consideration (Panel B), heritability and genetic correlations of the final and average model respectively (Panel C), and the effect size distribution of SNPs reported from the GWAS summary statistics for the paired traits (Panel D).

**Table 4:** Bidirectional estimated effect of differential WBC on schizophrenia estimated by MRCI method (β-coefficient are interpreted in units of standard deviation increase of absolute differential WBC count and causal effects at the liability scale of schizophrenia)

| Exposure | Outcome | Beta | P-value | FDR adjusted p-value |
| --- | --- | --- | --- | --- |
| Schizophrenia | Basophil count | -5.1e-3 | 0.985 | 1.000 |
| Schizophrenia | Eosinophil count | -7.1e-4 | 0.976 | 1.000 |
| Schizophrenia | Lymphocyte count | -2.2e-5 | 0.999 | 1.000 |
| Schizophrenia | Monocyte count | -1.2e-6 | 1.000 | 1.000 |
| Schizophrenia | Neutrophil count | -1.6e-5 | 0.999 | 1.000 |
| Basophil count | Schizophrenia | 0.021 | 0.642 | 0.642 |
| Eosinophil count | Schizophrenia | 0.048 | 0.010** | 0.026** |
| Lymphocyte count | Schizophrenia | 0.077 | 0.010** | 0.026** |
| Monocyte count | Schizophrenia | 0.032 | 0.061* | 0.102 |
| Neutrophil count | Schizophrenia | 0.027 | 0.318 | 0.398 |

\*p-value<0.1 , \*\*p-value<0.05

## 4. Discussion

We conducted bidirectional two-sample MR analyses using the two largest genomic datasets to-date on schizophrenia and WBC and investigated the possible causal relationships between the two traits, as well as putative causal mechanisms linking them. Our study used hundreds of SNPs to examine the bidirectional relationships, and the sample size was sufficient to detect true effects as low as 0.05 standard deviation away from the mean of differential WBC. In light of the substantial heterogeneity and outliers in the datasets, we carried out outlier-robust MR methods as well as the MRCI which explicitly modelled the situation of horizontal pleiotropy.

Our findings generally supported a causal relationship between raised lymphocyte count and schizophrenia. Based on the unbiased results generated from the MRCI approach and the results corrected for multiple testing, our analysis indicated that excess eosinophil count, which may disrupt the immune microenvironment, could contribute to causing schizophrenia. Furthermore, despite the possibilities of spurious estimates caused by pleiotropy from conventional MR approaches based on the substantial heterogeneity (Supplementary Table S2), the contamination mixture method suggested schizophrenia could possibly lead to reduction of eosinophil count and elevated neutrophil count. Such findings of schizophrenia reducing eosinophil count may implicate altered innate immune system in schizophrenia patients, and may partially explain the vulnerability to infection in schizophrenia patients (Pankiewicz-Dulacz et al., 2018). Moreover, elevated neutrophil count in schizophrenia has been reported in numerous studies on NLR (Karageorgiou et al., 2019), but causality could not be confirmed in the observational study designs. Our smaller effect sizes than the reported effect sizes in observational studies indicate that both environmental factors and genetic factors likely play roles in influencing the differential WBC, as the SNPs included in this study failed to explain all the differences.

Our findings are broadly consistent with the extant literature. For example, our finding supported the macrophage-T-lymphocyte theory of schizophrenia, i.e., activated T-lymphocytes play an important role in the neurotransmitter abnormalities and neuro-inflammation model of schizophrenia (Smith & Maes, 1995). Dopamine receptors were found to be expressed in different T cells and B cells, which could activate T cell functions in the human body (Levite, 2015). Activated T cells may then affect the dopamine activity in the brain, and therefore increase the risk of developing psychosis (Kesby et al., 2018). Besides, blood-borne monocytes were associated with the immune system in the central nervous system. Impaired and dysregulated monocyte activation were reported in schizophrenia (Uranova at el., 2017). Neutrophils are recognized as significant non-neural neurotransmitters that modulate central and peripheral nervous systems with crucial yet lesser known roles in pathophysiology of infectious, inflammatory, and neurological disorders (Kanashiro et al., 2020). Moreover, hyper-eosinophilia has long been linked to various neurological and psychiatric symptoms (Sin & Lee; 2014). Taken together, understanding the relationship between schizophrenia and different WBC may bring further insights in the relationships between schizophrenia and autoimmune diseases.

Another notable finding from our study was that a single SNP, rs13195636, appeared to be highly influential and pleiotropic in conventional MR methods after clumping. This SNP is located at the major histocompatibility complex (MHC) on chromosome 6. The population minor allele frequencies ranged from 1.8% in African to 10% in Non-Finnish North-West European (Open Targets, 2023). It is the strongest SNP reported by the PGC-schizophrenia and also highly significant in differential WBC reported by the Blood Cell Consortium. Genes located at MHC are believed to have critical roles in neurodevelopment and immune regulation (Gibney & Drexhage, 2013; Sekar et al., 2016). Closest protein coding gene of rs13195636 is ZNF184 gene, which is associated with bipolar disorder (Huang et al., 2022), depression (Wu et al., 2021), and Parkinson’s disease (Li et al., 2021). We speculated two possible explanations for this SNP. First, it could be owing to horizontal pleiotropy, which implies that this SNP has independent effects on multiple traits, including schizophrenia and different white blood cell subtypes. Another plausible explanation is that the effect size could possibly be inflated because of linkage disequilibrium with other genetic variants, which refers to non-random associations of alleles at multiple loci.

This study has several strengths. First, using the two largest publicly available genomic datasets of schizophrenia and differential WBC with hundreds of SNPs that had reached GWAS significance threshold, confers sufficient statistical power to detect small effects in this study. Although entering many instruments in the model is prone to weak instrument bias, this bias tends towards the null in two-sample MR (Yang et al., 2022). Therefore, significant results in this study were unlikely to be false positive due to weak instrument bias. Second, MR examined the causal relationships that typically could not be assessed using observational study design. Present of confounding factors, such as antipsychotic exposures, and potential reverse causation between immunological biomarkers and aetiology of schizophrenia, may bias the outcome measurements in observational research (Zhou et al., 2020; Dawidowski et al., 2022). Our two-sample MR study minimized the influence of confounders by including one sample with schizophrenia only and another sample with minimal subjects with schizophrenia. Third, we employed different MR methods, and carefully identified any violation against the MR assumptions. The novel MRCI method enabled us to obtain unbiased estimates with conservative standard errors. Lastly, we corrected the potential inflation of Type I error due to multiple testing using false discovery rate adjusted p-values.

Several limitations should be borne in mind. Although WBC to some extent shows trait-like properties, these indices are known to vary with time, even within a single day (Hilderink et al., 2017). This temporal variation can induce error and undermine the variance accounted for by the SNPs included in this study. Second, only autosome GWAS data of the Blood Cell Consortium were publicly available. Therefore, we only used autosome GWAS data from Psychiatric Genomics Consortium, and the potential influences of sex chromosomes (i.e., the widely-recognized gender differences in schizophrenia and WBC) were not captured in our results. In addition, using GWAS results from large meta-analysis studies may lead to higher risk of sample overlap between the exposure and outcome, which can bias the results to the null or the observational estimate (Hemani et al., 2018). Our results could be biased by a small portion of schizophrenia patients recruited into UK biobank who could not be identified and excluded from and the Blood Cell Consortium. Lastly, due to the different population structure of PGC and Blood Cell Consortium, our analysis only used European ancestry for fair comparison.

## 5. Conclusion

To conclude, the relationship between schizophrenia and the immunological system is complex. Our two-sample MR on two large datasets provided preliminary evidence that higher eosinophil and lymphocyte count, which are essential immune response biomarkers, could be causal susceptibility factors for developing schizophrenia; whereas schizophrenia could also increase neutrophil count and reduce eosinophil count. Future research should explore the immune-related biological mechanisms for schizophrenia, as well as the generalizability of our results in different ancestry and broader populations.

## Data Availability

All data produced are available online at Psychiatric Genomic Consortium and IEU Open GWAS Project

## 6. Acknowledgements

This study was funded by HKU Seed Fund for Basic Research for New Staff (202009185071) and the HKU Enhanced Start-up Fund for New Staff granted to Simon SY Lui.

The authors declare no competing interests.

