## Supplementary material for "Bidirectional two-sample Mendelian randomization study of differential white blood cell count and schizophrenia"

**Supplemental Materials - Contents**

### **Supplementary Table 1. Cochran’s Q and I2 statistics on different set of instruments used in MR methods of schizophrenia on differential WBC**

| **Exposure** | **Outcome** | **SNPs selected by method** | **Number of SNPs** | **F-statistics** | **Cochran’s Q** | **I^2^** |
| --- | --- | --- | --- | --- | --- | --- |
| Schizophrenia | Basophil count | GWAS significant threshold (p-value <=5e-8) | 146 | 44.42 | 489 | 0.978 |
|  |  | MR PRESSO | 139 | 43.54 | 234 | 0.977 |
|  |  | Contamination Mixture | 110 | 43.75 | 65 | 0.978 |
| Schizophrenia | Eosinophil count | GWAS significant threshold (p-value <=5e-8) | 146 | 44.42 | 1171 | 0.978 |
|  |  | MR PRESSO | 129 | 42.91 | 246 | 0.977 |
|  |  | Contamination Mixture | 102 | 42.58 | 81 | 0.977 |
| Schizophrenia | Lymphocyte count | GWAS significant threshold (p-value <=5e-8) | 146 | 44.42 | 1347 | 0.978 |
|  |  | MR PRESSO | 131 | 42.44 | 310 | 0.977 |
|  |  | Contamination Mixture | 93 | 42.62 | 78 | 0.977 |
| Schizophrenia | Monocyte count | GWAS significant threshold (p-value <=5e-8) | 146 | 44.42 | 1055 | 0.978 |
|  |  | MR PRESSO | 129 | 43.30 | 241 | 0.977 |
|  |  | Contamination Mixture | 99 | 43.36 | 73 | 0.978 |
| Schizophrenia | Neutrophil count | GWAS significant threshold (p-value <=5e-8) | 146 | 44.42 | 1087 | 0.978 |
|  |  | MR PRESSO | 122 | 43.58 | 223 | 0.977 |
|  |  | Contamination Mixture | 99 | 43.55 | 80 | 0.978 |

### **Supplementary Table 2. Cochran’s Q and I2 statistics on different set of instruments used in MR methods of differential WBC on schizophrenia**

| **Exposure** | **Outcome** | **SNPs selected by method** | **Number of SNPs** | **F-statistics** | **Cochran’s Q** | **I^2^** |
| --- | --- | --- | --- | --- | --- | --- |
| Basophil count | Schizophrenia | GWAS significant threshold (p-value <=5e-8) | 193 | 90.58 | 538 | 0.990 |
|  |  | MR PRESSO | 186 | 89.48 | 351 | 0.989 |
|  |  | Contamination Mixture | 137 | 99.17 | 94 | 0.990 |
| Eosinophil count | Schizophrenia | GWAS significant threshold (p-value <=5e-8) | 428 | 126.47 | 1089 | 0.992 |
|  |  | MR PRESSO | 417 | 124.95 | 834 | 0.992 |
|  |  | Contamination Mixture | 306 | 134.18 | 204 | 0.993 |
| Lymphocyte count | Schizophrenia | GWAS significant threshold (p-value <=5e-8) | 484 | 112.42 | 1384 | 0.991 |
|  |  | MR PRESSO | 462 | 108.58 | 930 | 0.991 |
|  |  | Contamination Mixture | 336 | 116.09 | 216 | 0.991 |
| Monocyte count | Schizophrenia | GWAS significant threshold (p-value <=5e-8) | 492 | 167.48 | 1422 | 0.994 |
|  |  | MR PRESSO | 470 | 168.61 | 961 | 0.994 |
|  |  | Contamination Mixture | 351 | 190.40 | 250 | 0.995 |
| Neutrophil count | Schizophrenia | GWAS significant threshold (p-value <=5e-8) | 412 | 102.64 | 1285 | 0.990 |
|  |  | MR PRESSO | 399 | 103.35 | 898 | 0.990 |
|  |  | Contamination Mixture | 282 | 112.22 | 217 | 0.991 |

### **Supplementary Figure 1. Single SNP effects of differential white blood cell count on schizophrenia**

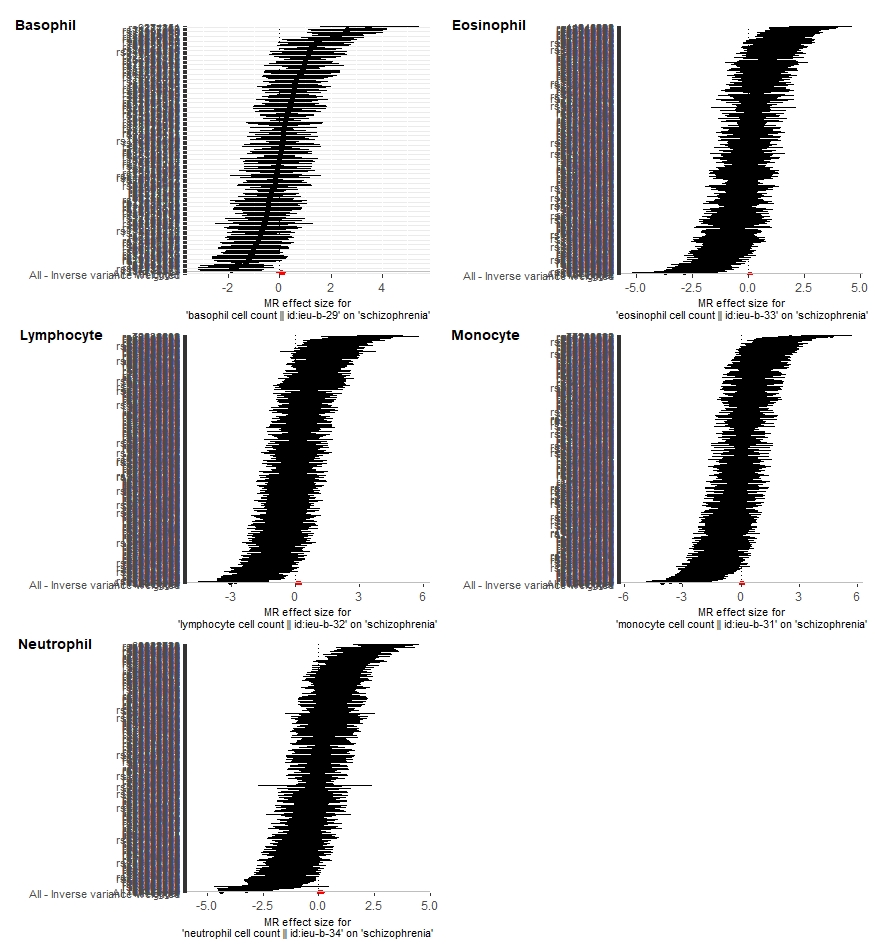

### **Supplementary Figure 2. Leave-one-out effects of differential white blood cell count on schizophrenia**

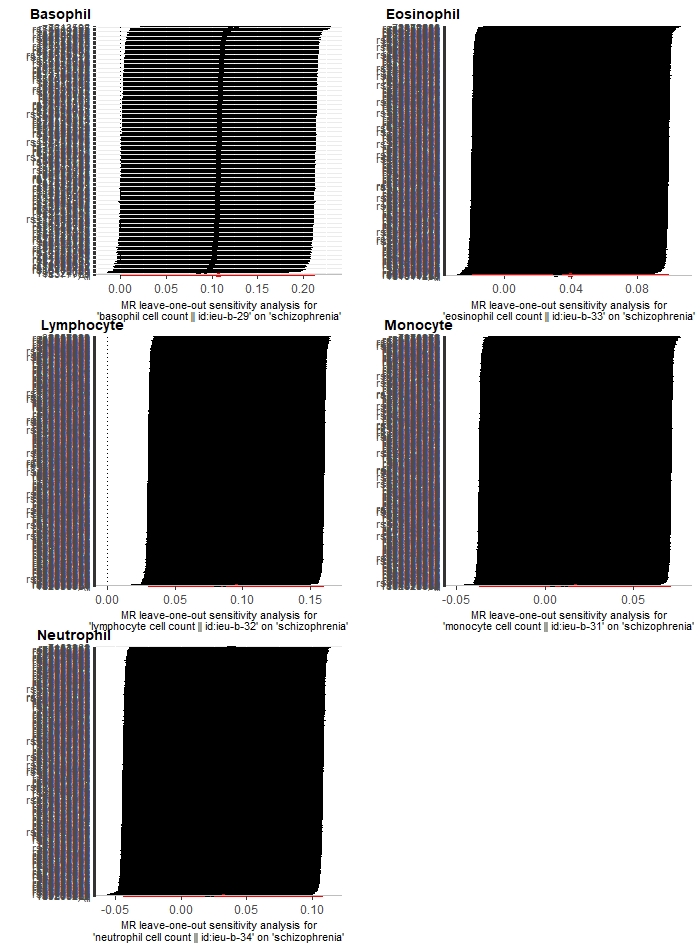

### **Supplementary Figure 3. Scatter plot of differential white blood cell count on schizophrenia**

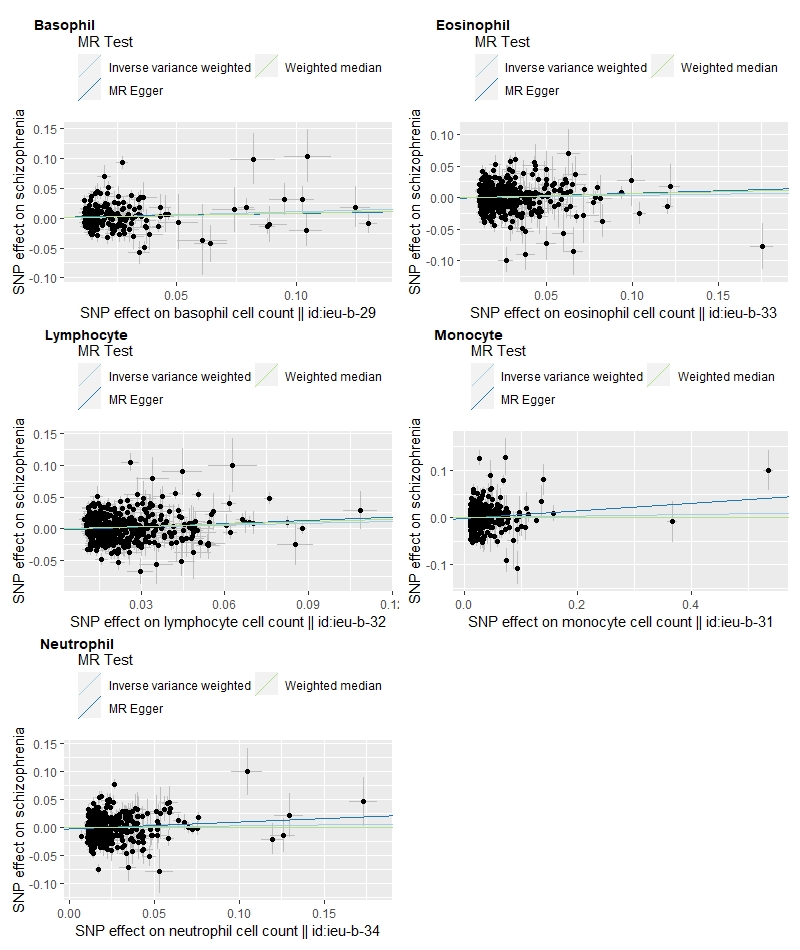

### **Supplementary Figure 4. Funnel plots of differential white blood cell count on schizophrenia**

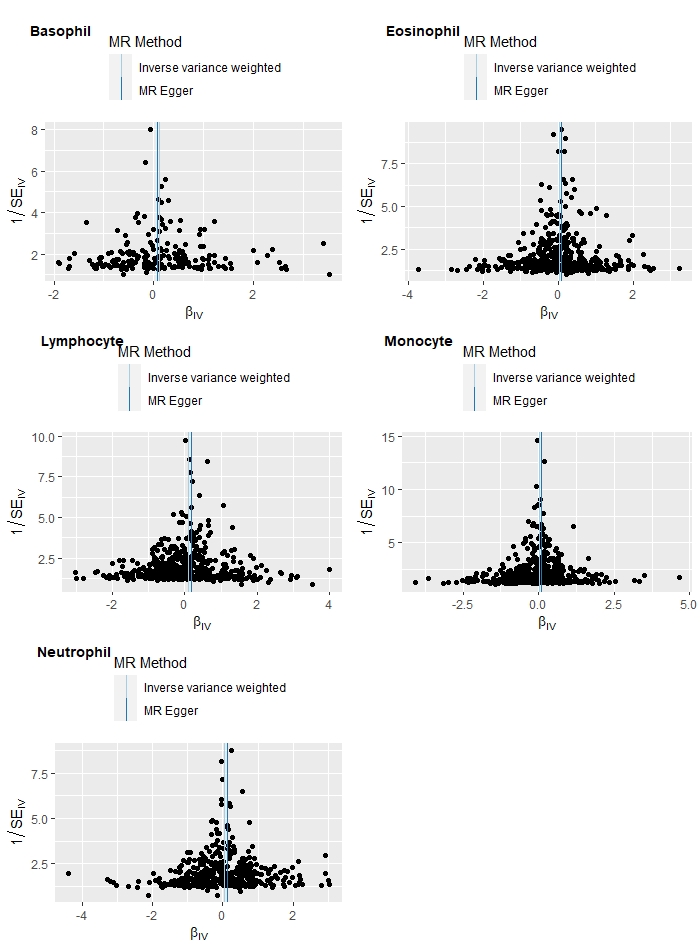

### **Supplementary Figure 5. Single SNP effects of schizophrenia on differential white blood cell count**

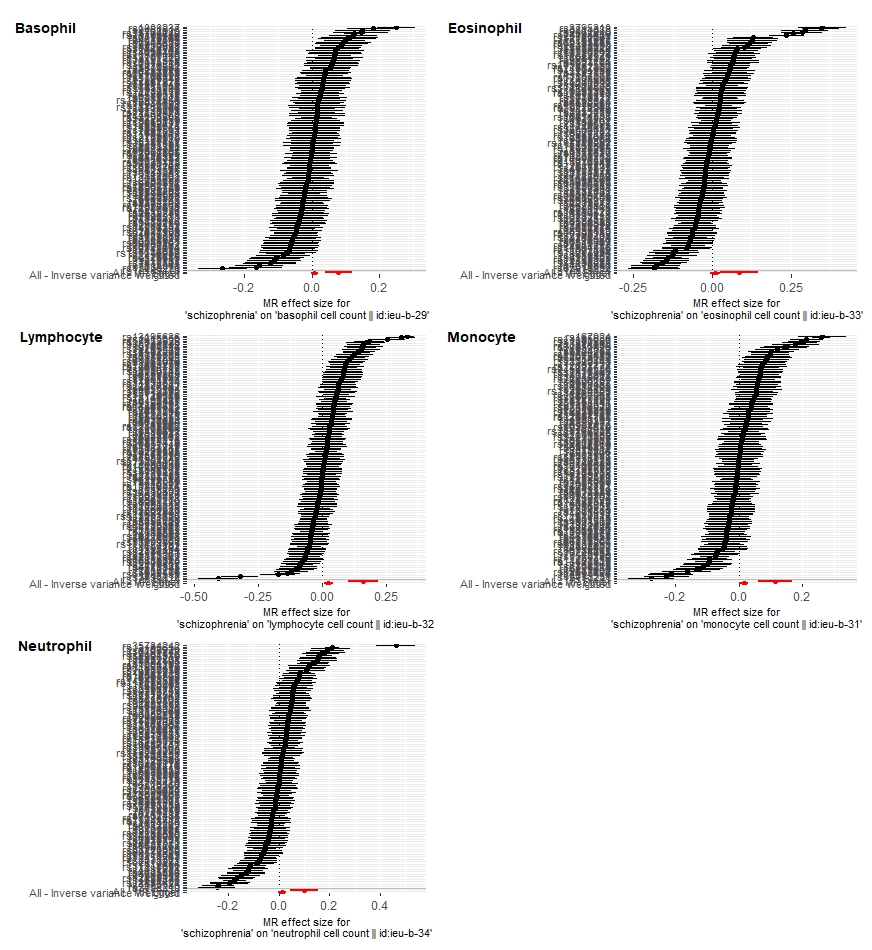

### **Supplementary Figure 6. Leave-one-out effects of schizophrenia on differential white blood cell count**

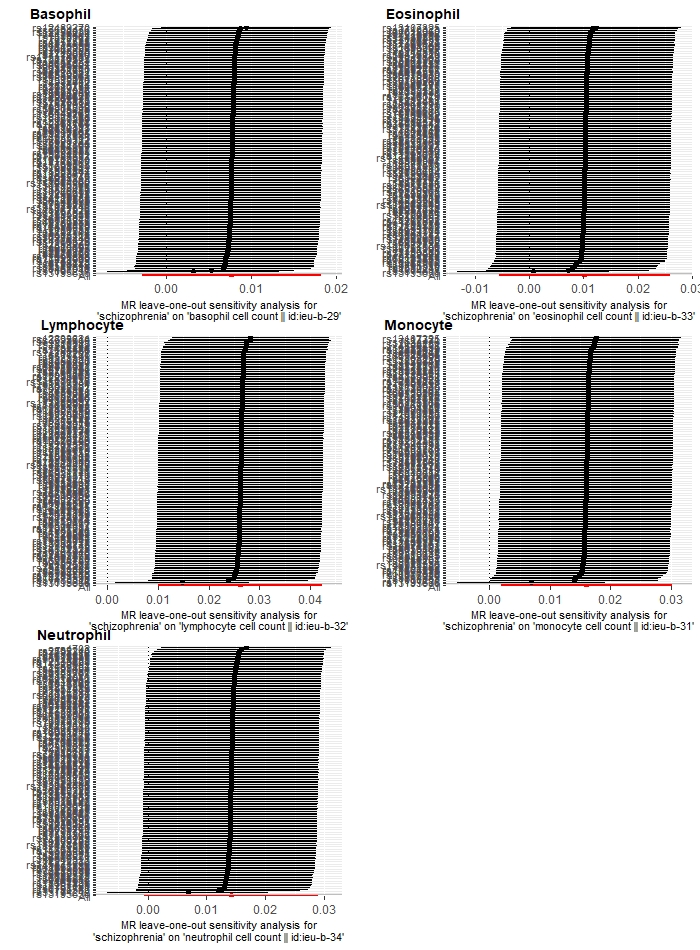

### **Supplementary Figure 7. Scatter plot of schizophrenia on differential white blood cell count**

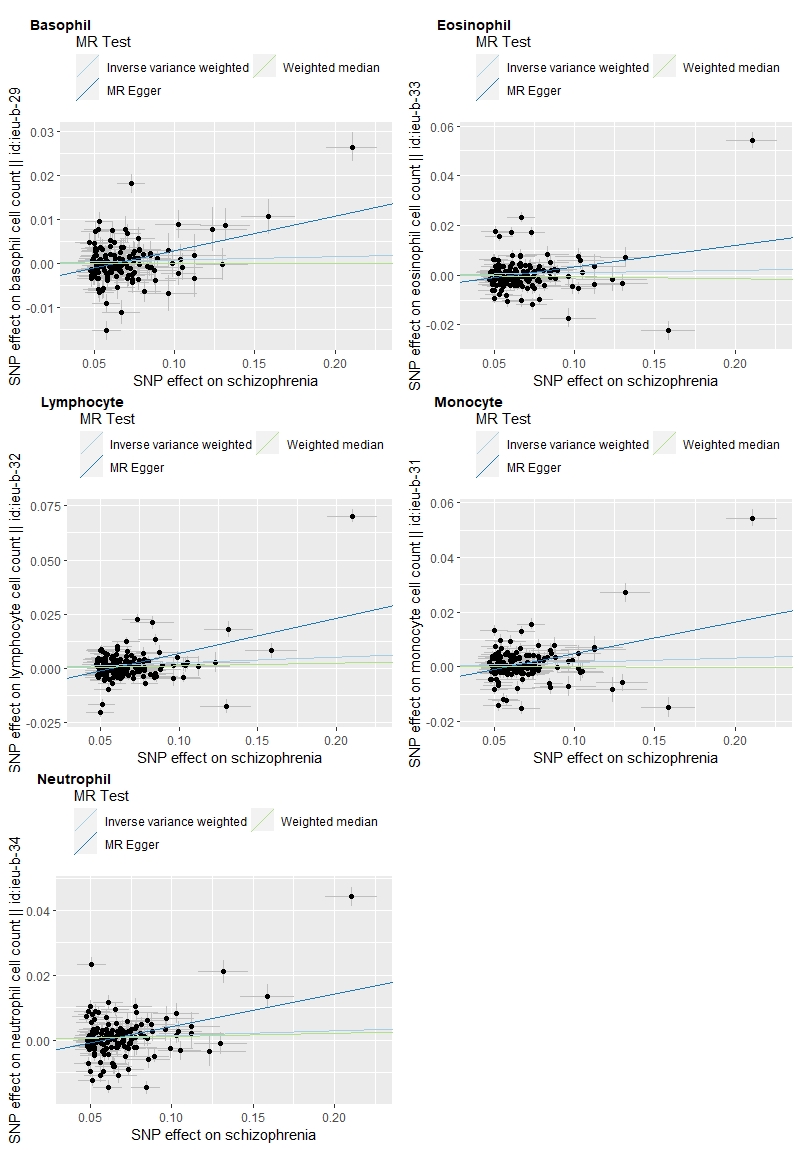

### **Supplementary Figure 8. Funnel plots of schizophrenia on differential white blood cell count**

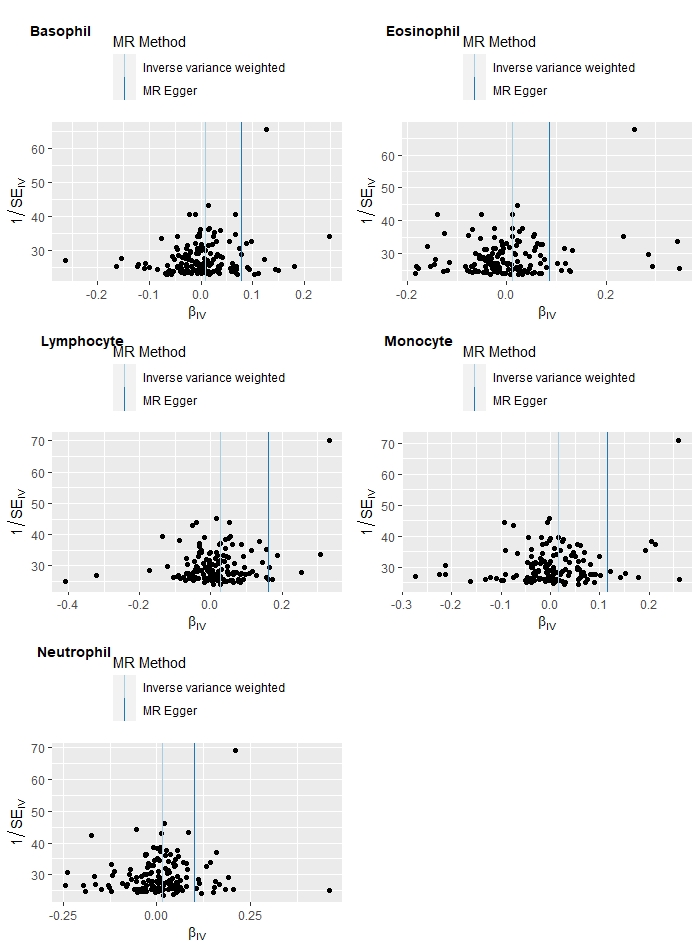

### **Supplementary Figure 9. Summary results of MRCI on schizophrenia and basophil count**

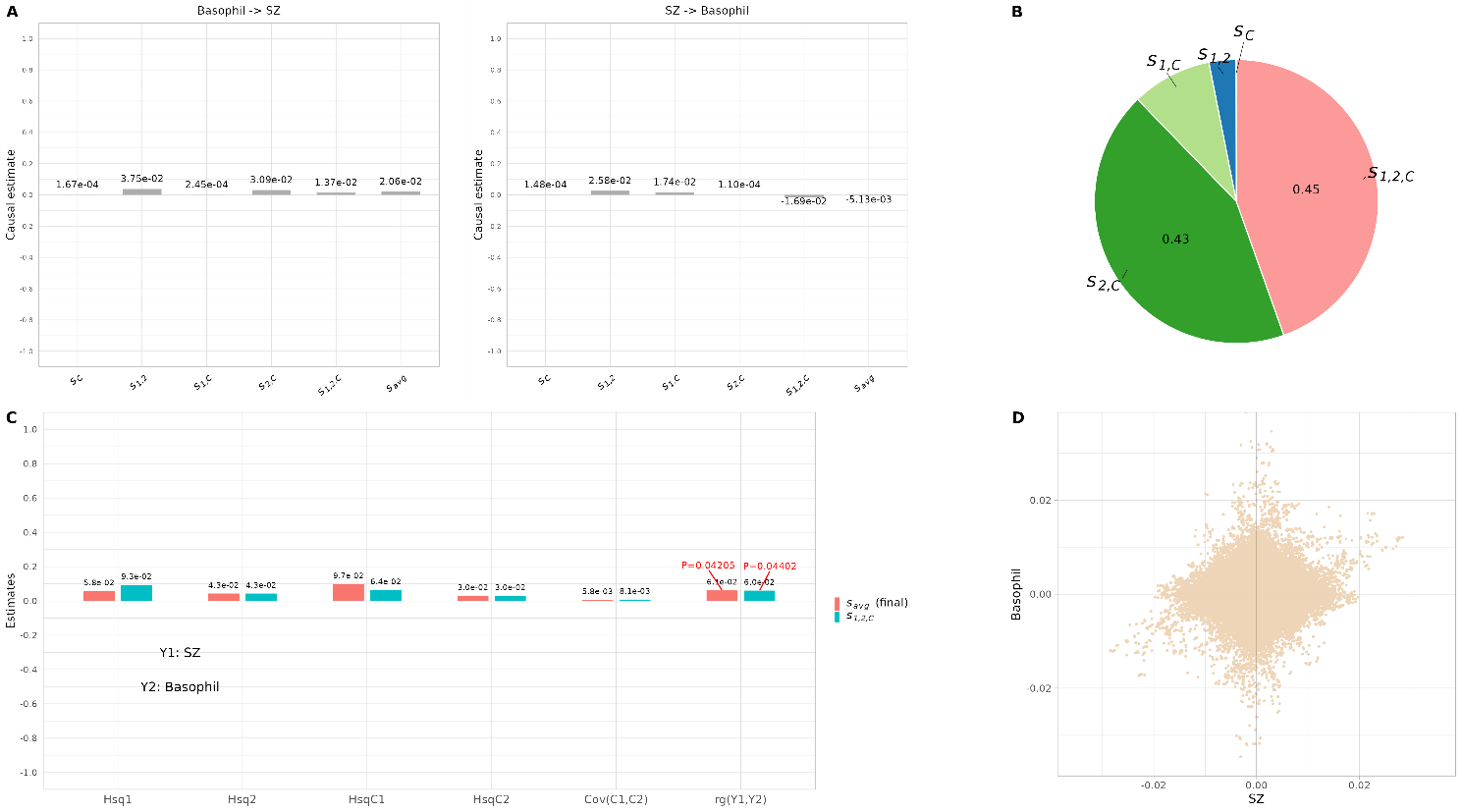

### **Supplementary Figure 10. Summary results of MRCI on schizophrenia and eosinophil count**

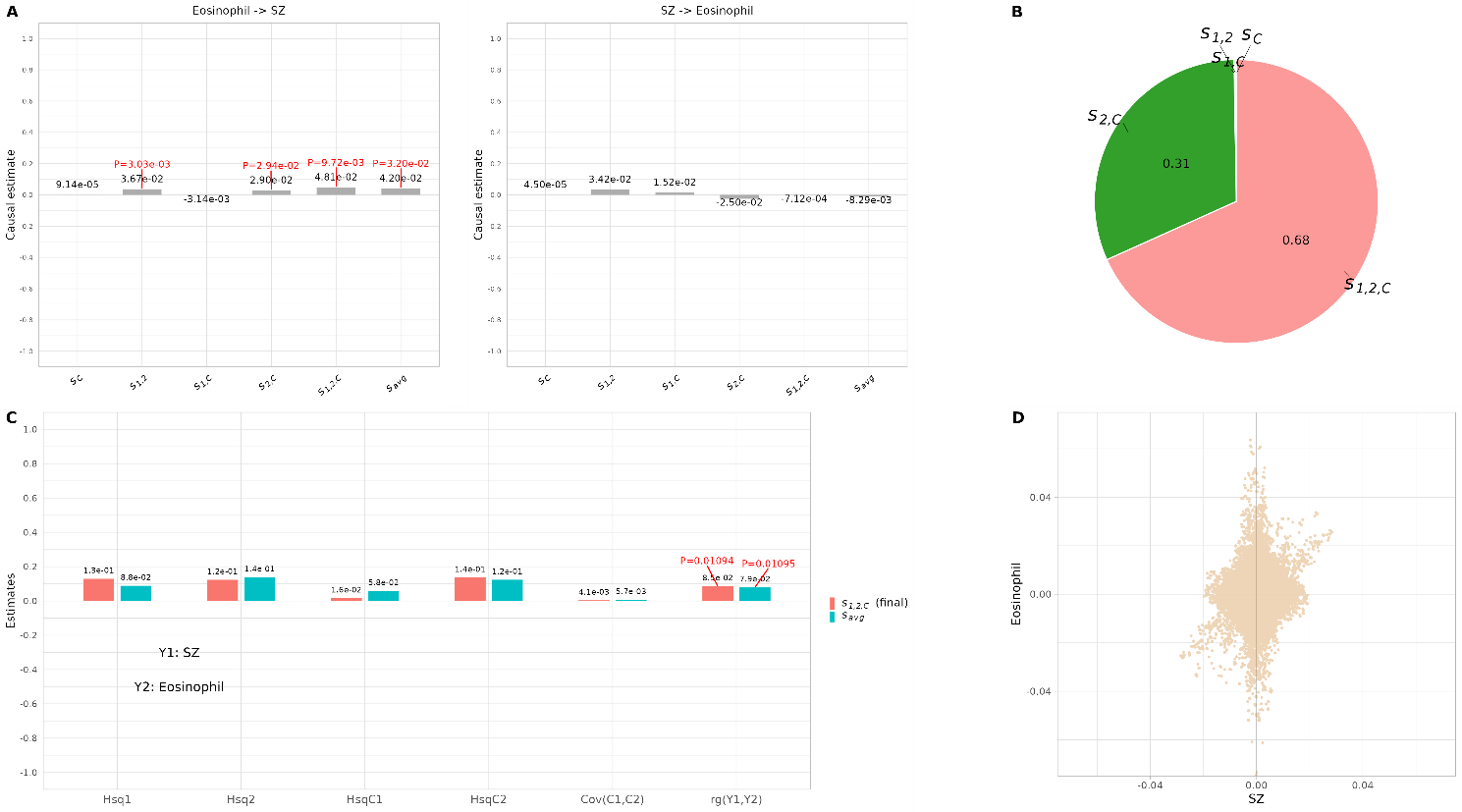

### **Supplementary Figure 11. Summary results of MRCI on schizophrenia and lymphocyte count**

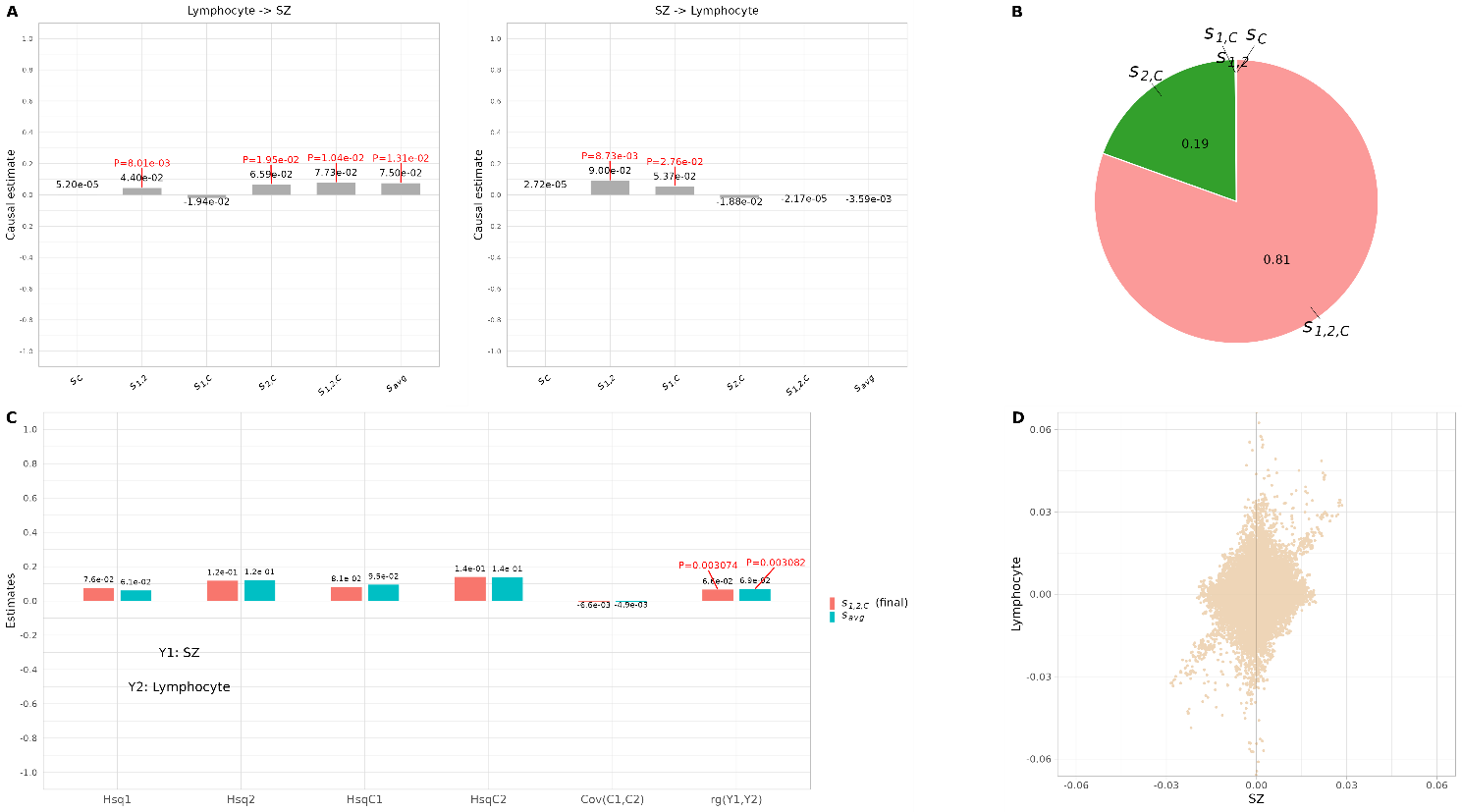

### **Supplementary Figure 12. Summary results of MRCI on schizophrenia and monocyte count**

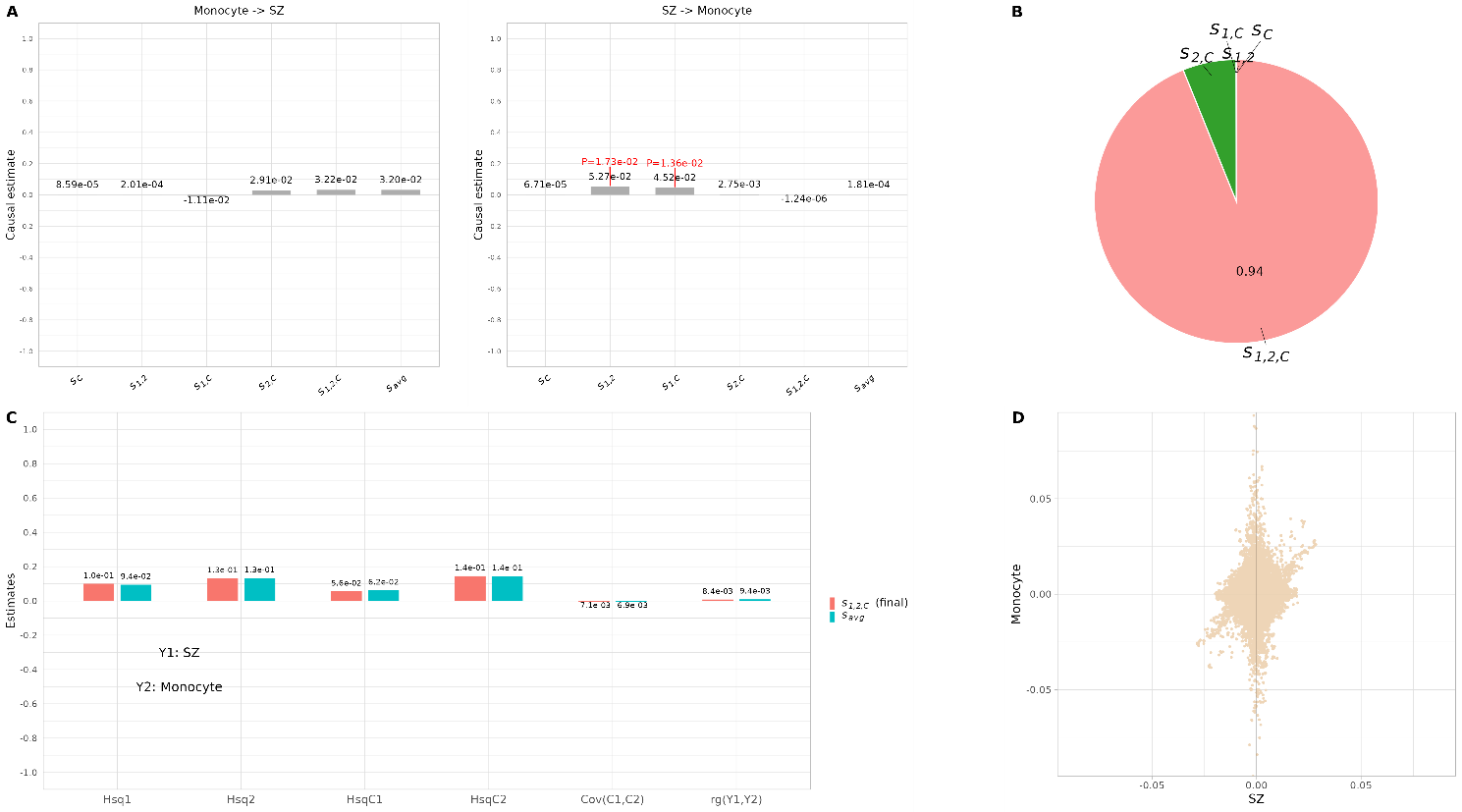

### **Supplementary Figure 13. Summary results of MRCI on schizophrenia and neutrophil count**

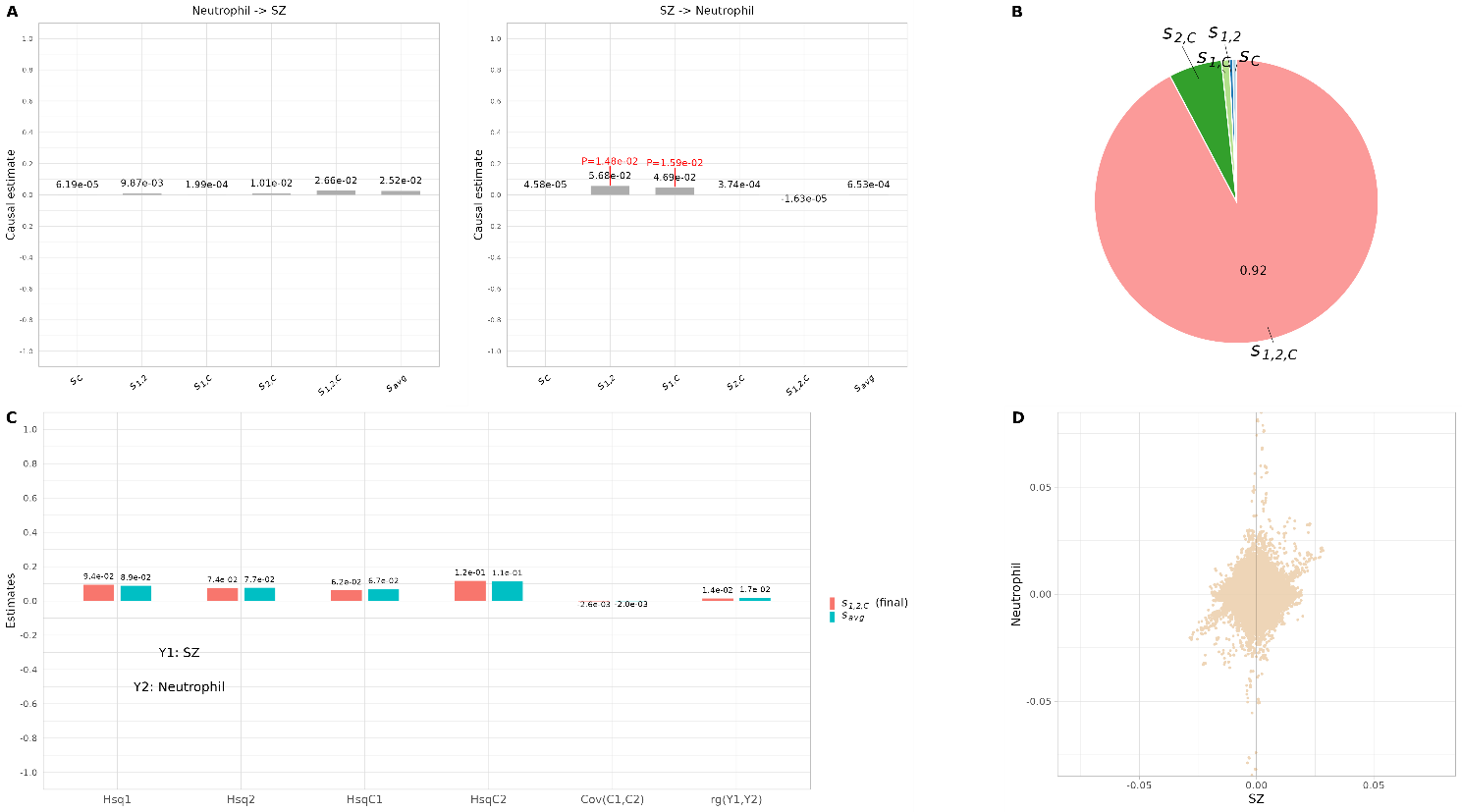
